# Temperature-Driven Variability in Emergency Diagnostic Accuracy by a Leading Language Model

**DOI:** 10.1101/2025.06.04.25328288

**Authors:** Philip C. Jarrett, Jared Hill, Marshall Howell, Kristen Grabow Moore, Joby J. Thoppil, Laura Vargas Ortiz, Samuel T. Parnell, D. Mark Courtney, Samuel A. McDonald, Deborah B. Diercks, Andrew R. Jamieson, Dazhe Cao

**Affiliations:** Department of Emergency Medicine, The University of Texas Southwestern Medical Center, Dallas, TX, USA; Department of Emergency Medicine, Emory University School of Medicine, Atlanta, GA, USA; Lyda Hill Department of Bioinformatics, The University of Texas Southwestern Medical Center, Dallas, TX, USA

**Keywords:** Artificial Intelligence, Diagnosis, Computer-Assisted, Emergency Service, Hospital, Clinical Decision Support Systems

## Abstract

**Objective:** To determine the impact of the temperature parameter on GPT-4o’s diagnostic accuracy when evaluating emergency medicine cases and assess the effect on diagnostic divergence across iterations.

**Methods:** We conducted a simulation-based diagnostic accuracy study using four challenging emergency medicine cases adapted from the Foundations of Emergency Medicine curriculum. Each case was submitted to GPT-4o 250 times at five temperature settings (0.0, 0.25, 0.50, 0.75, 1.0), both with and without physical examination findings, yielding 10,000 total outputs. Each output contained exactly three differential diagnoses with one leading diagnosis. Diagnostic accuracy was assessed by comparing outputs against predetermined gold-standard diagnoses.

**Results:** At temperature 0.0, GPT-4o achieved 100% leading diagnosis accuracy across all cases with physical exam data. As temperature increased, accuracy declined systematically to 89.4% at temperature 1.0. Diagnostic divergence increased dramatically from an average of 4.5 unique diagnoses at temperature 0.0 to 26.25 at temperature 1.0 (583% increase). Case sensitivity varied significantly, with ascending cholangitis showing the greatest temperature sensitivity (accuracy dropping from 100% to 70.4%) while carbon monoxide poisoning maintained 100% accuracy across all settings.

**Discussion:** Higher temperatures introduced concerning diagnostic inconsistency rather than beneficial exploration, with substantial accuracy degradation in temperature-sensitive cases.

**Conclusions:** Lower temperature settings promote diagnostic accuracy and consistency, making them preferable for clinical applications requiring high reliability. Transparent reporting of temperature settings is essential for reproducible clinical artificial intelligence research.

**KEY MESSAGES:** *What is already known on this topic:* Large language models demonstrate promising diagnostic capabilities in medical reasoning tasks, but their non-deterministic nature and sensitivity to parameter settings remain poorly understood in clinical contexts.

*What this study adds:* The temperature parameter significantly affects both diagnostic accuracy and consistency, with higher settings causing dramatic increases in diagnostic divergence.

*How this study might affect research, practice or policy:* These findings mandate transparent reporting of temperature settings in clinical AI research and suggest that low-temperature configurations should be prioritized for high-reliability medical applications.

## INTRODUCTION

Large language models (LLMs) such as OpenAI’s GPT-4o have demonstrated significant medical reasoning capabilities, with studies suggesting diagnostic accuracy comparable to physicians [1–6]. These models are being applied to clinical decision support, particularly in high-stakes environments like emergency medicine (EM) [7–12]. LLMs are also integral to ambient dictation solutions that formulate clinical notes on behalf of physicians [13]. However, LLM performance is non-deterministic; it can vary with repetitions of the same task and can be influenced by various parameters [14–18].

Temperature, in the context of LLMs, is a parameter that controls the randomness of a model’s output [14–18]. A low temperature setting (e.g., 0.0 or 0.2) consistently leads to focused, predictable outputs. Conversely, a high temperature (e.g., 0.8 or 1.0) increases randomness, allowing the model to sample less likely responses. This can result in more diverse outputs, but also increases the risk of inappropriate responses to clinical tasks.

Temperature may impact the reliability of LLM-generated medical guidance [17,18]. If an LLM suggests differential diagnoses in clinical notes, a low temperature should offer the most probable diagnoses, potentially missing rarer conditions. A high temperature might suggest more creative possibilities but could also introduce "noise" in the form of improbable diagnoses. The choice of temperature setting represents a trade-off between consistency and exploration.

Despite its importance, temperature settings are underreported in LLM clinical research, limiting reproducibility [1–3,16–19]. Additionally, much of the literature on LLM diagnostic performance has focused on challenging models with rare conditions, misrepresenting daily practice [1–3]. Furthermore, the default temperature settings in commercial chatbots are often unknown to the user, eroding trust in outputs intended for clinical use [14]. There is a need to systematically evaluate how temperature affects LLM performance on medical tasks, particularly in scenarios that mimic real-world data limitations, such as early patient encounters in the emergency department where only initial history of present illness (HPI), vitals, and perhaps physical exam data are available.

The primary objective of this study was to determine the impact of the temperature parameter on GPT-4o’s diagnostic accuracy when evaluating high-risk EM cases based on early clinical information. Secondary objectives included assessing the effect of temperature on the diagnostic divergence generated and understanding the consistency of model outputs across a range of temperatures.

## METHODS

### Study Design and Case Selection

We conducted a simulation-based diagnostic accuracy study using GPT-4o (Figure 1). Four challenging EM cases were adapted from the Foundations of Emergency Medicine (FoEM) curriculum, a widely adopted training resource protected by login, minimizing the likelihood of inclusion in GPT-4o’s training data [20–21]. Case selection was guided by several criteria, as determined by editors of the FoEM curriculum. Each case must lead to a single unambiguous diagnosis. The clinical information must represent a cross-sectional snapshot of an early patient encounter (chief complaint, vitals, HPI, past medical/surgical history, and physical examination). The cases must reflect diagnoses spanning a range of organ systems.

**Figure 1.**
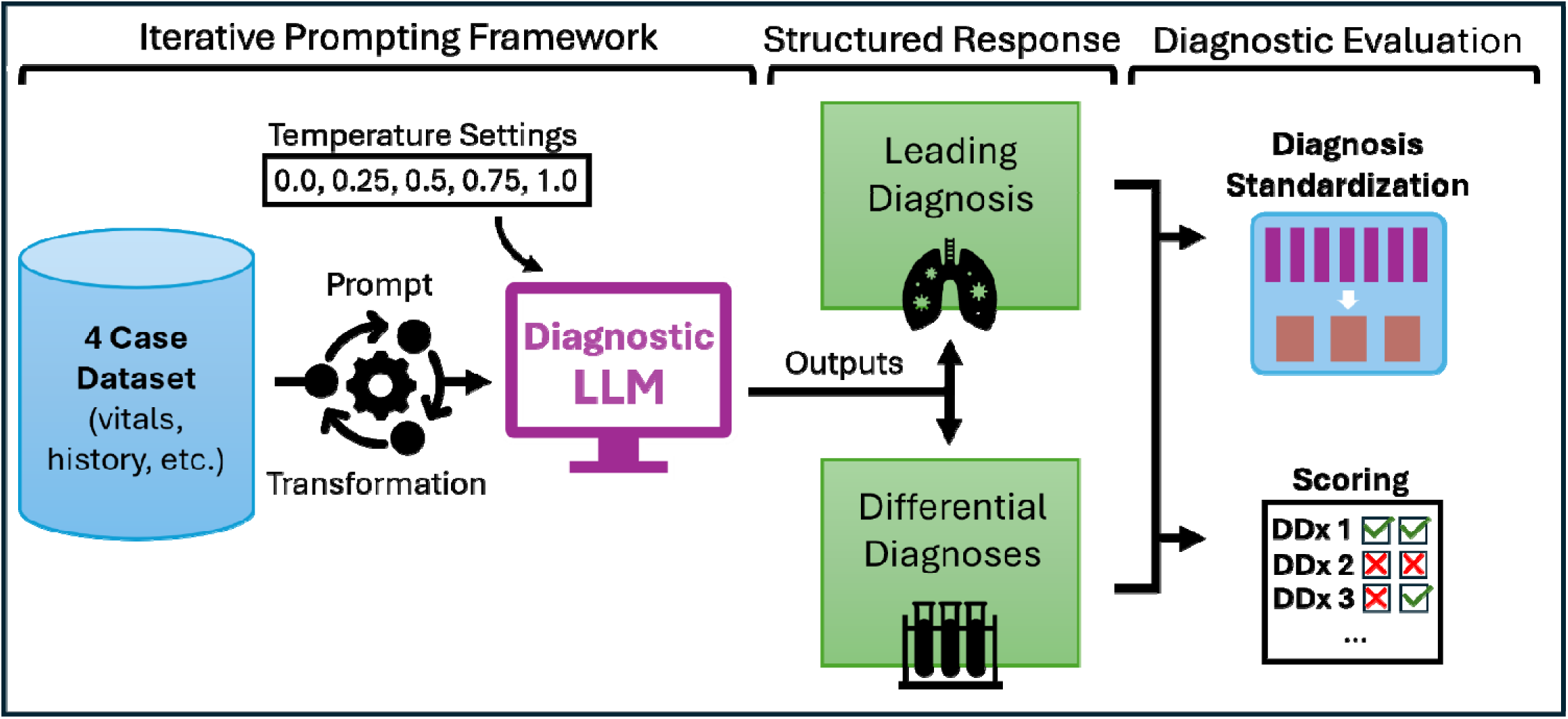
Study flow chart demonstrating the diagnostic evaluation pipeline using a large language model (LLM) and output scoring. Structured cases from Foundations of Emergency Medicine were adapted to represent early-encounter patient information. A standardized prompt specified the diagnostic reasoning task and output structure. The diagnostic LLM (GPT-4o) returned structured responses. Diagnostic lists (DDx) were scored for accuracy while unique diagnoses were standardized to assess diagnostic breadth.

**Figure 2.**
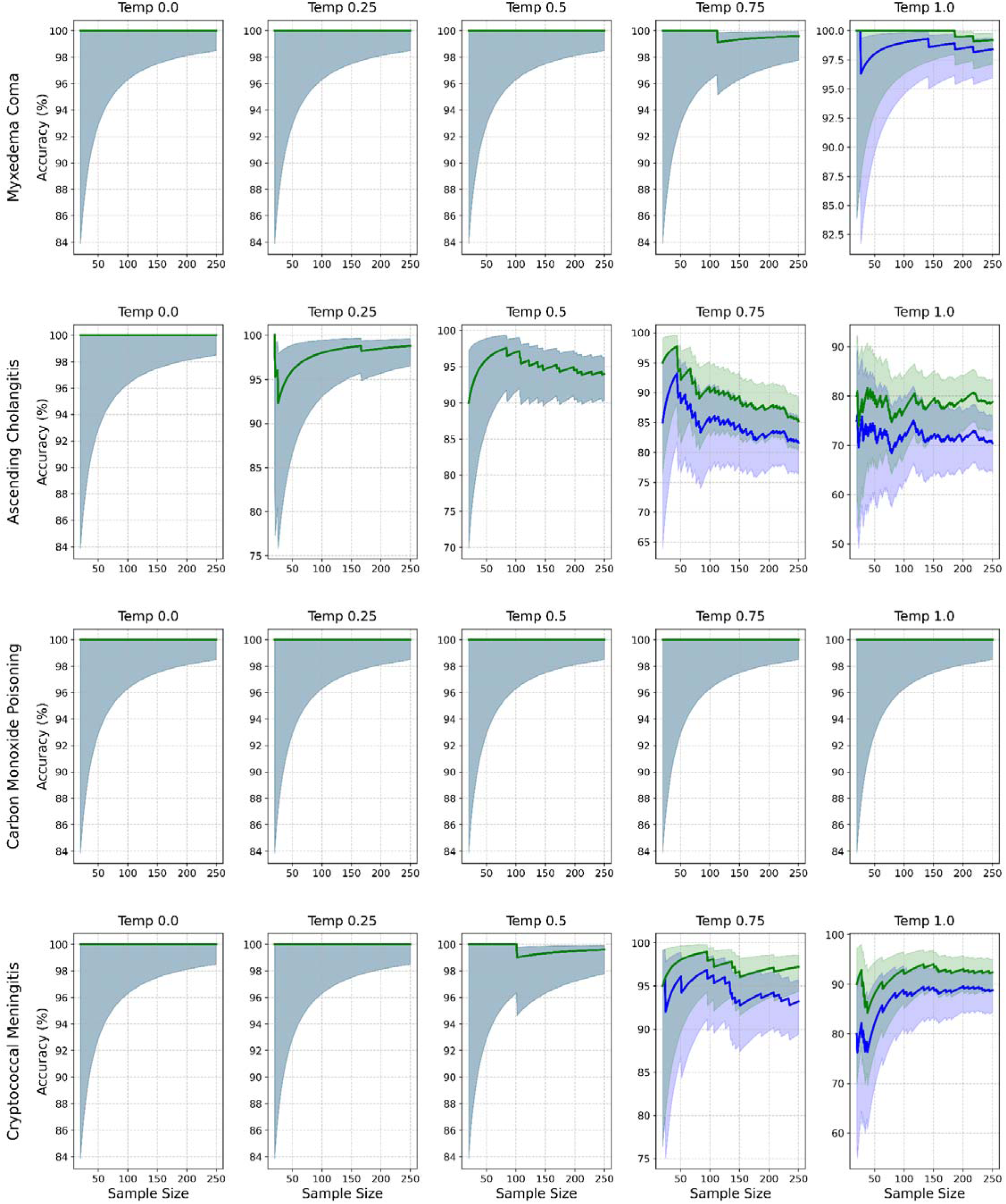
Diagnostic accuracy by case (rows) and temperature (columns) with exams. Lines show accuracy for leading (blue) and differential (green) diagnoses. Shaded pink and yellow areas represent the 95% con idence interval (CI) for the leading and differential diagnoses, respectively. Grey areas represent overlap in 95% CI.

The diagnoses associated with these cases served as the gold standard for assessing diagnostic accuracy (Table 1). The selected cases were Myxedema Coma (Endocrinology), Ascending Cholangitis (Infectious Disease/Gastroenterology), Carbon Monoxide Poisoning (Toxicology), and Cryptococcal Meningitis (Neurology/Infectious Disease).

**Table 1.**
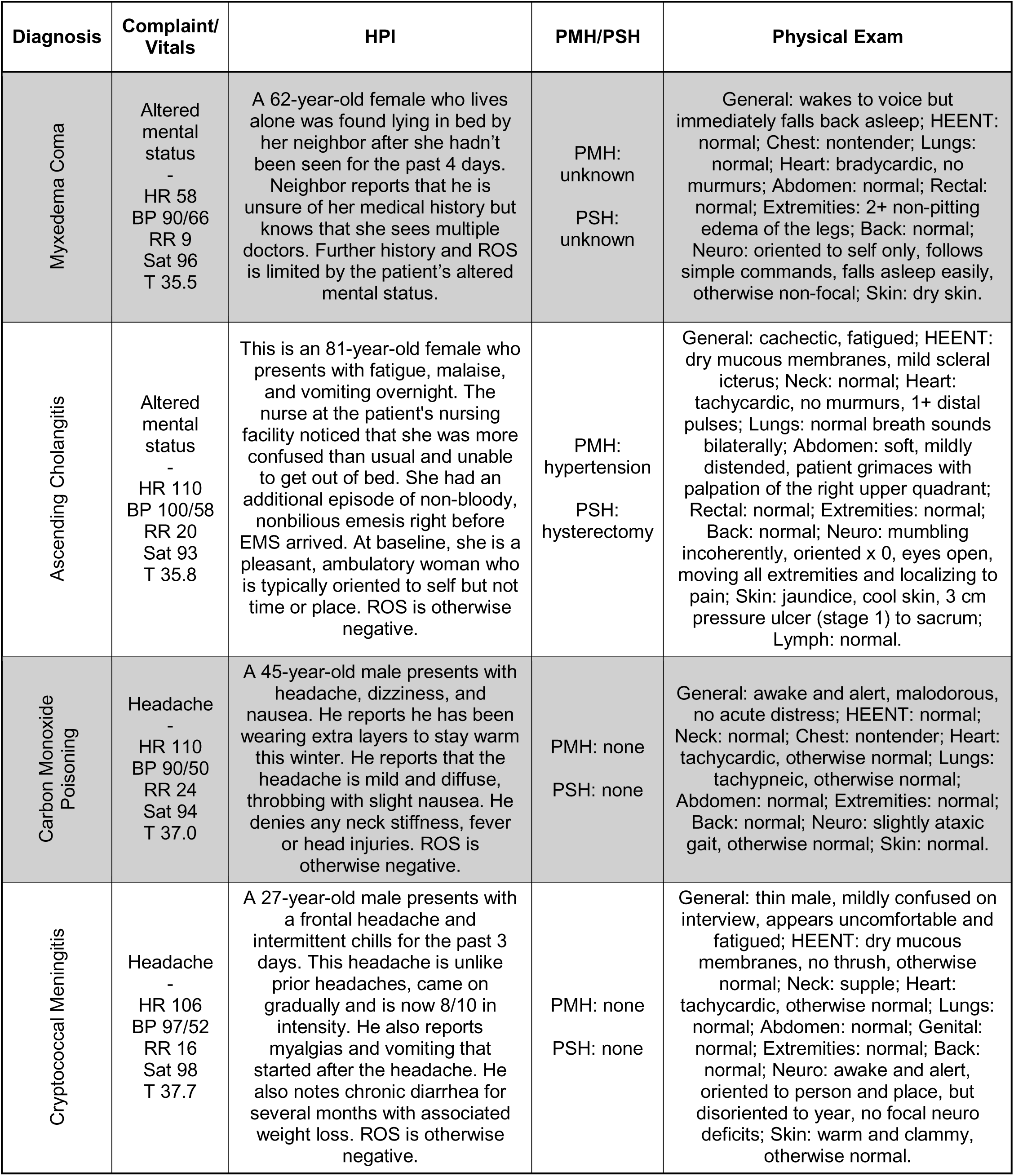
Details of the four cases selected from the Foundations of Emergency Medicine curriculum to evaluate GPT-4o diagnostic accuracy. HPI: history of present illness. PMH: past medical history. PSH: past surgical history. HR: heart rate. BP: blood pressure. RR: respiratory rate. Sat: oxygen saturation. T: body temperature. HEENT: head, eyes, ears, nose, throat. ROS: review of systems.

| Diagnosis | Complaint/<br>Vitals | HPI | PMH/PSH | Physical Exam |
| --- | --- | --- | --- | --- |
| Myxedema Coma | Altered mental status<br>-<br>HR 58<br>BP 90/66<br>RR 9<br>Sat 96<br>T 35.5 | A 62-year-old female who lives alone was found lying in bed by her neighbor after she hadn't been seen for the past 4 days. Neighbor reports that he is unsure of her medical history but knows that she sees multiple doctors. Further history and ROS is limited by the patient's altered mental status. | PMH: unknown<br>PSH: unknown | General: wakes to voice but immediately falls back asleep; HEENT: normal; Chest: nontender; Lungs: normal; Heart: bradycardic, no murmurs; Abdomen: normal; Rectal: normal; Extremities: 2+ non-pitting edema of the legs; Back: normal; Neuro: oriented to self only, follows simple commands, falls asleep easily, otherwise non-focal; Skin: dry skin. |
| Ascending Cholangitis | Altered mental status<br>-<br>HR 110<br>BP 100/58<br>RR 20<br>Sat 93<br>T 35.8 | This is an 81-year-old female who presents with fatigue, malaise, and vomiting overnight. The nurse at the patient's nursing facility noticed that she was more confused than usual and unable to get out of bed. She had an additional episode of non-bloody, nonbilious emesis right before EMS arrived. At baseline, she is a pleasant, ambulatory woman who is typically oriented to self but not time or place. ROS is otherwise negative. | PMH: hypertension<br>PSH: hysterectomy | General: cachectic, fatigued; HEENT: dry mucous membranes, mild scleral icterus; Neck: normal; Heart: tachycardic, no murmurs, 1+ distal pulses; Lungs: normal breath sounds bilaterally; Abdomen: soft, mildly distended, patient grimaces with palpation of the right upper quadrant; Rectal: normal; Extremities: normal; Back: normal; Neuro: mumbling incoherently, oriented x 0, eyes open, moving all extremities and localizing to pain; Skin: jaundice, cool skin, 3 cm pressure ulcer (stage 1) to sacrum; Lymph: normal. |
| Carbon Monoxide Poisoning | Headache<br>-<br>HR 110<br>BP 90/50<br>RR 24<br>Sat 94<br>T 37.0 | A 45-year-old male presents with headache, dizziness, and nausea. He reports he has been wearing extra layers to stay warm this winter. He reports that the headache is mild and diffuse, throbbing with slight nausea. He denies any neck stiffness, fever or head injuries. ROS is otherwise negative. | PMH: none<br>PSH: none | General: awake and alert, malodorous, no acute distress; HEENT: normal; Neck: normal; Chest: nontender; Heart: tachycardic, otherwise normal; Lungs: tachypneic, otherwise normal; Abdomen: normal; Extremities: normal; Back: normal; Neuro: slightly ataxic gait, otherwise normal; Skin: normal. |
| Cryptococcal Meningitis | Headache<br>-<br>HR 106<br>BP 97/52<br>RR 16<br>Sat 98<br>T 37.7 | A 27-year-old male presents with a frontal headache and intermittent chills for the past 3 days. This headache is unlike prior headaches, came on gradually and is now 8/10 in intensity. He also reports myalgias and vomiting that started after the headache. He also notes chronic diarrhea for several months with associated weight loss. ROS is otherwise negative. | PMH: none<br>PSH: none | General: thin male, mildly confused on interview, appears uncomfortable and fatigued; HEENT: dry mucous membranes, no thrush, otherwise normal; Neck: supple; Heart: tachycardic, otherwise normal; Lungs: normal; Abdomen: normal; Genital: normal; Extremities: normal; Back: normal; Neuro: awake and alert, oriented to person and place, but disoriented to year, no focal neuro deficits; Skin: warm and clammy, otherwise normal. |

Because physical exam data is often unavailable during early clinical encounters, we conducted a sensitivity analysis excluding physical exam to assess impact on diagnostic performance.

### Language Model Selection

GPT-4o (model version ‘gpt-4o-2024-05-13’ via the OpenAI Application Programming Interface) was chosen as the LLM for this study due to its widespread availability and strong performance in early medical benchmarks [14].

### Iterative Sampling and Prompting

Temperature parameters typically spans a range of 0.0 (low randomness) – 2.0 (high randomness). Each of the four cases was iterated 250 times at five distinct temperature settings (0.0, 0.25, 0.50, 0.75, and 1.0), both with and without physical exam findings. Temperatures above 1.0 were excluded because preliminary testing indicated impaired adherence to prompt instructions at higher values. Iterative prompting across all case-temperature pairs resulted in 10,000 total outputs.

All prompting followed a structured format designed to simulate consultation with an artificially intelligent (AI) diagnostic assistant (Supplement 1). To standardize evaluation, the model was restricted to producing exactly three differential diagnoses per case, including one leading diagnosis. This constraint served several purposes. First, limiting output length reduced the risk of inflated diagnostic accuracy that can occur when evaluating correctness within long, unranked lists of conditions. This issue has been previously noted as a pattern of overtesting by LLMs, maximizing sensitivity at the cost of specificity [22]. Second, it better reflects real-world diagnostic decision-making, where clinicians apply heuristics to prioritize clinical possibilities when determining workup and treatment. Third, a three-item limit helped isolate the model’s prioritization behavior, forcing it to make judgment calls about what diagnoses warrant inclusion under constraint. This structure also enabled assessment of diagnostic divergence across iterations, by observing variability in top-ranked differentials across multiple runs.

Each prompt began with a standardized header, which included instructions about the diagnostic task, expected output formatting, and clinical information about a single case (chief complaint, vital signs, history of present illness, past medical history, and — if applicable — physical exam findings). In the sensitivity analysis excluding physical exam data, this section was replaced with the phrase "unavailable at this time." Prompts were designed to be stateless and were submitted in parallel, ensuring that the model had no memory of prior case iterations. Additionally, internet search and external reference capabilities were disabled during all interactions, requiring GPT-4o to rely exclusively on its embedded knowledge to generate diagnostic responses.

### Outcome Measures

The primary outcomes for this study were specified a priori. Diagnostic Accuracy refers to the correctness of the diagnoses proposed by GPT-4o when compared against a pre-specified, definitive diagnosis for each case, known as the "gold standard." Diagnostic accuracy was assessed in two ways: Leading Diagnosis (whether the single diagnosis listed by GPT-4o as its first and most likely diagnosis matched the gold standard diagnosis for that case) and Differential Diagnosis (whether the gold standard diagnosis for that case appeared anywhere within the three-item list of differential diagnoses provided by GPT-4o).

Diagnostic Divergence was measured by the total number of unique diagnoses proposed by the LLM for each case, tallied at each temperature setting. This metric quantifies inconsistency in diagnostic outputs across iterations with identical inputs, representing variability in the differential diagnosis across case repetitions.

### Diagnostic Accuracy Scoring

A keyword search was used to score diagnostic accuracy for each of the 10,000 outputs. For each case, specific keyword phrases representative of the correct diagnosis were predefined: Myxedema Coma ("myxedema"), Ascending Cholangitis ("cholangitis"), Carbon Monoxide Poisoning ("carbon monoxide", "co"), and Cryptococcal Meningitis ("cryptococcal meningitis", "cryptococcal meningoencephalitis", "cryptococcal encephalitis").

The listed diagnoses (leading and three-item differential) provided by GPT-4o were searched for these keywords (case-insensitive). If a keyword was found in the leading diagnosis, it was scored as correct for "Leading Diagnosis". If found in any of the three differential diagnoses, it was scored as correct for "Differential Diagnosis". Diagnostic accuracy with and without inclusion of physical exam are plotted in Figure 3 and Supplement 2, demonstrating the 95% confidence intervals for diagnostic performance through iterative sampling.

To validate this keyword scoring method, two medical students (JH, LVO), blinded to the keyword scores and temperature settings, independently reviewed 1,000 outputs (250 from each case, sampled evenly across all temperatures and physical exam inclusion states). The reviewers scored the leading and differential diagnoses as correct or incorrect based on the gold standard diagnosis for each case. Disagreements between the two student reviewers were adjudicated by a board-certified EM physician (PJ). Following adjudication, there was perfect concordance between the keyword search and human review (Cohen’s κ = 1.0), confirming the reliability of this systematic scoring method.

### Diagnostic Divergence Analysis

Each individual LLM output was constrained to a three-item differential diagnosis, representing the model’s top-priority conditions based on the presented case. However, when the same case is presented repeatedly, the model may generate different sets of diagnoses each time due to the non-deterministic nature of LLMs. This across-iteration variability — despite identical input — represents diagnostic inconsistency that undermines reliability and suggests randomness exaggerated by adjustment of the temperature parameter.

To quantify this inconsistency, we measured diagnostic divergence as the total number of unique diagnoses generated across all iterations of a given case at each temperature setting (Figure 4). The relative frequencies of each diagnosis across case iterations are provided in Supplement 3. To isolate the role of temperature in this analysis, only cases that included physical exam data were included (n = 5,000). Because diagnoses were often expressed with subtle differences in terminology, all outputs were standardized to group synonymous or conceptually equivalent terms. This allowed us to calculate the total number of distinct, standardized diagnostic categories per case-temperature pair.

**Figure 4.**
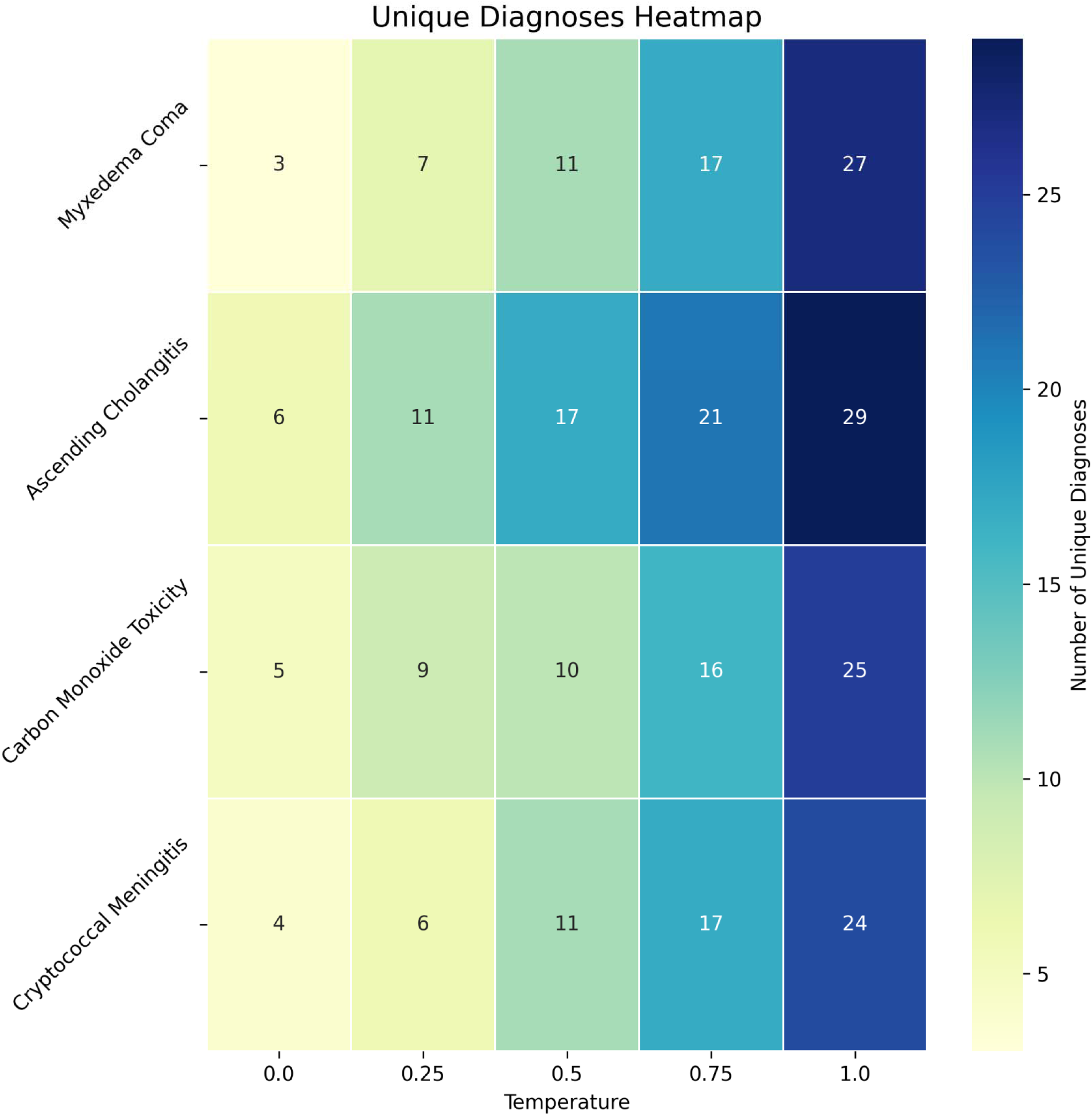
Heatmap depicting the number of standardized diagnoses considered in the three-item differential diagnoses across repetitive samples for each case-temperature pair. Diagnoses may overlap, such that items counted at low temperatures may also be counted at higher temperatures.

### Statistical Analysis

Diagnostic accuracy (leading and differential) was calculated as a percentage with corresponding 95% confidence intervals (CIs) using the Wilson score interval method. These metrics were calculated for each case-temperature pair. The number of unique diagnoses (diagnostic divergence) was tabulated. All analyses were performed using Python (Version 3.9).

### Ethics and Reporting

This study was approved by the Institutional Review Board of the author’s academic institution during expedited review of a parent protocol. The present study does not involve human subjects. The authors adhered to the TRIPOD-LLM Reporting Guidelines for studies using or evaluating LLMs.

## RESULTS

### Diagnostic Scoring Procedure

The keyword search was applied to all 10,000 LLM outputs to score leading and differential diagnoses as correct or incorrect. To validate this approach, a random 10% sample (n=1,000) was independently reviewed by two medical students who were successfully blinded to the scores of the keyword search function. Reviewer agreement was high, with only three discrepancies between humans across all 1,000 cases (Cohen’s κ = 0.99). These three cases were escalated to a board-certified emergency physician, also blinded to the keyword-based scores. In all instances, the tiebreak aligned with the keyword search. Following adjudication, there was perfect concordance between the keyword search and human review (Cohen’s κ = 1.0), confirming the reliability of this systematic scoring method.

### Diagnostic Accuracy

When provided physical exam data, GPT-4o demonstrated high diagnostic accuracy, particularly at lower temperature settings, achieving 100% accuracy for the leading diagnosis at temperature 0.0 across all cases.

As temperature increased, a clear trend of decreasing accuracy was observed (Table 2). Overall leading diagnosis accuracy declined from 100% at temperature 0.0 to 89.4% at temperature 1.0 (95% CI: 85.8 - 91.8). Accuracy of the three-item differential diagnosis remained higher but showed a similar decline, dropping from 100% at lower temperatures to 92.6% at temperature 1.0 (95% CI: 89.3 – 94.6).

**Table 2.** Diagnostic accuracy (95% confidence intervals [CI]) of GPT-4o on each case at five pre-specified temperatures including and excluding physical exam findings.

| Case | Temp | Accuracy (+exams) |  | Accuracy (-exams) |  |
| --- | --- | --- | --- | --- | --- |
|  |  | Leading | Differential | Leading | Differential |
| Myxedema Coma | 0.0 | 100% (98.5-100) | 100% (98.5-100) | 0% (0-1.5) | 100% (98.5-100) |
|  | 0.25 | 100% (98.5-100) | 100% (98.5-100) | 0% (0-1.5) | 96% (92.8-97.8) |
|  | 0.5 | 100% (98.5-100) | 100% (98.5-100) | 3.6% (1.9-6.7) | 93.2% (89.4-95.7) |
|  | 0.75 | 99.6% (97.8-99.9) | 99.6% (97.8-99.9) | 3.6% (1.9-6.7) | 89.2% (84.7-92.5) |
|  | 1.0 | 98.4% (96-99.4) | 99.2% (97.1-99.8) | 9.2% (6.2-13.4) | 82.8% (77.6-87) |
| Ascending Cholangitis | 0.0 | 100% (98.5-100) | 100% (98.5-100) | 0% (0-1.5) | 0% (0-1.5) |
|  | 0.25 | 98.8% (96.5-99.6) | 98.8% (96.5-99.6) | 0% (0-1.5) | 0% (0-1.5) |
|  | 0.5 | 94% (90.3-96.3) | 94% (90.3-96.3) | 0% (0-1.5) | 0% (0-1.5) |
|  | 0.75 | 81.6% (76.3-85.9) | 85.2% (80.3-89.1) | 0% (0-1.5) | 0% (0-1.5) |
|  | 1.0 | 70.4% (64.5-75.7) | 78.8% (73.3-83.4) | 0% (0-1.5) | 0% (0-1.5) |
| Carbon Monoxide Toxicity | 0.0 | 100% (98.5-100) | 100% (98.5-100) | 100% (98.5-100) | 100% (98.5-100) |
|  | 0.25 | 100% (98.5-100) | 100% (98.5-100) | 100% (98.5-100) | 100% (98.5-100) |
|  | 0.5 | 100% (98.5-100) | 100% (98.5-100) | 100% (98.5-100) | 100% (98.5-100) |
|  | 0.75 | 100% (98.5-100) | 100% (98.5-100) | 100% (98.5-100) | 100% (98.5-100) |
|  | 1.0 | 100% (98.5-100) | 100% (98.5-100) | 100% (98.5-100) | 100% (98.5-100) |
| Cryptococcal Meningitis | 0.0 | 100% (98.5-100) | 100% (98.5-100) | 100% (98.5-100) | 100% (98.5-100) |
|  | 0.25 | 100% (98.5-100) | 100% (98.5-100) | 100% (98.5-100) | 100% (98.5-100) |
|  | 0.5 | 99.6% (97.8-99.9) | 99.6% (97.8-99.9) | 97.6% (94.9-98.9) | 97.6% (94.9-98.9) |
|  | 0.75 | 93.2% (89.4-95.7) | 97.2% (94.3-98.6) | 94.4% (90.8-96.6) | 95.6% (92.3-97.5) |
|  | 1.0 | 88.8% (84.3-92.1) | 92.4% (88.4-95.1) | 82.8% (77.6-87) | 85.6% (80.7-89.4) |
| Overall | All | 96.2% (95.7-96.7) | 97.2% (96.7-97.7) | 49.6% (48.2-50.9) | 72% (70.7-73.2) |

The impact of temperature varied significantly by case (Table 2). Ascending Cholangitis demonstrated the most sensitivity to temperature changes, with leading diagnosis accuracy dropping from 100% at temperature 0.0 to 70.4% at temperature 1.0 (95% CI: 64.5 – 75.7). In contrast, Carbon Monoxide Poisoning maintained 100% accuracy across all temperatures, likely due to a pathognomonic presentation. Myxedema Coma and Cryptococcal Meningitis showed intermediate sensitivity to temperature increases.

### Sensitivity Analysis of Physical Exam

Excluding physical exam data led to a substantial reduction in diagnostic performance overall, with accuracy for the leading diagnosis dropping to 49.6% (95% CI: 48.2 – 51.0) and the three-item differential to 72.0% (95% CI: 70.7 – 73.2). The effect varied by case. Myxedema Coma was particularly sensitive in terms of the leading diagnosis, which dropped to 3.3% (95% CI: 2.4 – 4.4), although the correct diagnosis still appeared within the differential list in 92.2% (95% CI: 90.6 – 93.6) of iterations. Ascending Cholangitis showed complete dependence on physical exam findings — diagnostic accuracy fell to 0% for both the leading and differential diagnoses in their absence. Carbon Monoxide Poisoning was unaffected by the removal of physical exam data, with diagnostic accuracy remaining at 100% across all temperatures. Cryptococcal Meningitis showed only minimal sensitivity to the physical exam, with leading diagnosis accuracy at 95.0% (95% CI: 93.6–96.0) and differential diagnosis accuracy at 95.8% (95% CI: 94.5–96.7).

### Diagnostic Divergence Analysis

To isolate the impact of temperature on diagnostic divergence, this analysis was limited to cases that included physical exam data. Across all case-temperature pairs, GPT-4o generated 371 unique raw diagnoses (case-insensitive). These were subsequently consolidated into 91 standardized diagnostic categories through human review.

Temperature had a significant effect on the inconsistency of diagnoses generated by GPT-4o across iterations (Figure 4). Increasing temperature consistently led to greater diagnostic divergence for each case. Myxedema Coma showed unique diagnoses increasing from 3 at temperature 0.0 to 27 at temperature 1.0. Ascending Cholangitis demonstrated unique diagnoses rising from 6 at temperature 0.0 to 29 at temperature 1.0. Carbon Monoxide Poisoning showed unique diagnoses increasing from 5 at temperature 0.0 to 25 at temperature 1.0. Cryptococcal Meningitis exhibited unique diagnoses growing from 4 at temperature 0.0 to 24 at temperature 1.0.

Overall, the average number of unique diagnoses was 4.5 at temperature 0.0, increasing to an average of 26.25 at temperature 1.0. This represents a 583% increase in diagnostic divergence across iterations of the same case. Higher temperatures led to inconsistent three-item differentials across case iterations.

As temperature increased to 1.0, the correct diagnosis often remained one of the most frequent, but its overall frequency typically decreased, and a larger set of alternative diagnoses appeared more often (Figure 5). For instance, in the case of ascending cholangitis at temperature 1.0, the correct diagnosis was still a common response, but other conditions appeared with increasing frequency, diluting diagnostic consensus across iterations.

## DISCUSSION

This study systematically evaluated the influence of temperature on GPT-4o’s diagnostic performance using challenging EM cases and data inputs typical of an early patient encounter. Our findings demonstrate that temperature significantly impacts both the accuracy and the consistency of diagnostic outputs generated by GPT-4o.

The primary observation is the trade-off mediated by temperature: lower temperatures (e.g., 0.0) yielded consistent and highly accurate diagnostic outputs. At these settings, GPT-4o reliably identified the correct diagnosis as its leading choice. This suggests that low-temperature settings constrain the model to prioritize high-probability conditions, which may be desirable in clinical workflows where precision and reliability are paramount.

Conversely, higher temperatures led to greater diagnostic divergence — manifesting as inconsistency across iterations. While this broader differential could superficially appear to reflect diagnostic breadth, it actually introduced problematic variability and degraded the accuracy of the leading diagnosis. For some cases, like ascending cholangitis, the decline in diagnostic accuracy at higher temperatures was substantial. These findings underscore a critical consideration for clinical use: in settings where LLMs are queried only once (e.g., through consumer-facing chatbots or synchronous clinical decision support tools), high temperature settings may introduce an unacceptable level of diagnostic uncertainty.

The clinical relevance of these findings extends to ongoing applications of LLMs in healthcare. In ambient documentation systems that generate clinical impressions and summarize patient encounters, low temperature settings may support accurate and consistent labeling of routine conditions — but could risk missing atypical diagnoses. However, users seeking broader differential diagnosis should utilize strategic prompting rather than increase temperature parameters, as higher temperatures primarily introduce inconsistency rather than systematically expanding diagnostic consideration. When higher temperatures are unavoidable, repeated sampling may be necessary to better characterize the distribution of potential diagnostic outputs, though this approach introduces additional cost and complexity.

Furthermore, temperature settings are typically hidden in commercial products, making it difficult for clinicians to judge the reliability of LLM-generated suggestions [14]. This lack of transparency has implications for trust, medicolegal accountability, and model governance. Even subtle variability in outputs can erode clinician confidence and create friction in environments where decisions must be made quickly, often with incomplete data.

Our findings offer guidance for developers and early adopters of LLM-driven tools in clinical settings. Minimizing temperature can support focused, reproducible outputs in documentation and clinical decision support contexts. When broader exploration is desired, structured iterative approaches may mitigate risk by identifying stable diagnostic signals across multiple model outputs.

Case-specific differences also emerged. Carbon monoxide poisoning maintained 100% accuracy across all temperatures, likely due to distinctive historical cues. In contrast, ascending cholangitis — whose symptoms overlap with many intra-abdominal processes — proved highly sensitive to temperature changes. These observations suggest that model performance under varying temperatures may not be uniform across diseases, and temperature tuning strategies may need to be context-specific.

Finally, our findings reinforce the importance of methodological transparency in LLM research. The temperature parameter significantly influenced diagnostic performance, yet it remains inconsistently reported in published studies. Without clarity around settings like temperature, top-p, and context length, readers cannot interpret or reproduce findings. Standardized reporting frameworks (e.g., TRIPOD-LLM) should be adopted broadly to improve interpretability and comparability across studies.

These findings have significant implications for the safe integration of LLMs into clinical workflows. Temperature tuning must be informed by the intended clinical use case: whether the goal is diagnostic triage, collaborative reasoning with human clinicians, or research applications. For users seeking broader differential diagnosis, prompt modification rather than temperature adjustment should be the preferred approach.

Transparent reporting of temperature settings—by both researchers and LLM vendors—is essential for ensuring reproducibility, interpretability, and trust. Ultimately, while LLMs like GPT-4o show considerable promise in medical reasoning, human oversight remains indispensable, particularly when interpreting outputs generated with higher degrees of randomness or when applied in safety-critical environments like the emergency department.

## LIMITATIONS

Simulated cases cannot replicate the full ambiguity of real presentations. Scoring relied on discrete gold-standard diagnoses rather than probabilistic reasoning. We restricted output length, which may favor precision. Only one LLM version was tested; generalizability to other models and future releases is unknown.

## CONCLUSIONS

Temperature is a decisive yet underreported lever in LLM-based diagnosis. Low settings optimize accuracy and reproducibility; high settings amplify diagnostic divergence and must be paired with iterative sampling or rigorous human oversight. Transparent reporting and thoughtful tuning are essential as LLMs enter safety-critical workflows like emergency medicine.

## Data Availability

All data produced in the present study are available upon reasonable request to the authors.

## Contributions

PJ, DC, and SP conceived of the research question and study design. PJ and DC contributed to the statistical methods and data analysis. MH and KM contributed to FoEM case selection and validation of a priori critical test selection. JH, LVO, and PJ contributed to blinded human review of critical test scoring. PJ, JH, MH, KM, JT, SM, SP, DD, MC, and DC contributed to writing the original draft, while MC, SM, LVO and AJ provided extensive editorial review. PJ, DC, SM, and AJ contributed to study supervision. The Cursor Integrated Development Environment was used for code development and execution using Anthropic’s Claude 3.5 Sonnet and Claude 3.7 Sonnet +/- Thinking. GPT-4o was used to support manuscript drafting after development and execution of all study methods. The authors thoroughly reviewed and revised all code and manuscript content. The lead author assumes all responsibility for the content of this manuscript. PJ takes responsibility for the paper as a whole.

## Acknowledgments

Many thanks are owed to Dr. Jeremy Berberian, Dr. Alison G. Marshall, and Dr. Rory Merritt for the original authorship of the FoEM cases used in this study, as well as to the numerous editors who peer-reviewed the cases for inclusion in the FoEM curriculum. We are also grateful to Katherine Riley Martin, M.S., for her expertise in the regulatory approval of this research.

---

**<u>Supplement 1.</u>** Diagnostic assistant prompt header used to iteratively collect responses from GPT-4o.The individualized case details were adjusted for each of the four cases. When physical exam data was excluded during sensitivity analysis, the exam details were replaced by the statement: “unavailable at this time.”

You are a Large Language Model (LLM) assisting with emergency department diagnostic evaluations. You will be provided with patient data (chief complaint, vitals, history of present illness (HPI), past medical/surgical history (PMH/PSH), and physical exam) collected during triage evaluation of the patient.

Your task is to produce exactly three differential diagnoses for the most likely underlying condition, including a leading diagnosis and two additional differential diagnoses. Diagnoses should be as specific as possible. Do not use imprecise terms such as delirium, sepsis or shock - such conditions should have an underlying source. A diagnosis of "Delirium secondary to urinary tract infection" should just be "urinary tract infection", representing the underlying cause. Do not use terms such as "possible" or "probable" in the diagnoses. Each diagnosis should be a single condition without qualifiers.

Chief Complaint: [Case-specific complaint]

Vitals: [Case-specific vitals]

History of Present Illness: [Case-specific HPI]

Past Medical/Surgical History: [Case-specific PMH/PSH]

Physical Examination: [Case-specific exam findings]

**Supplement 2.**
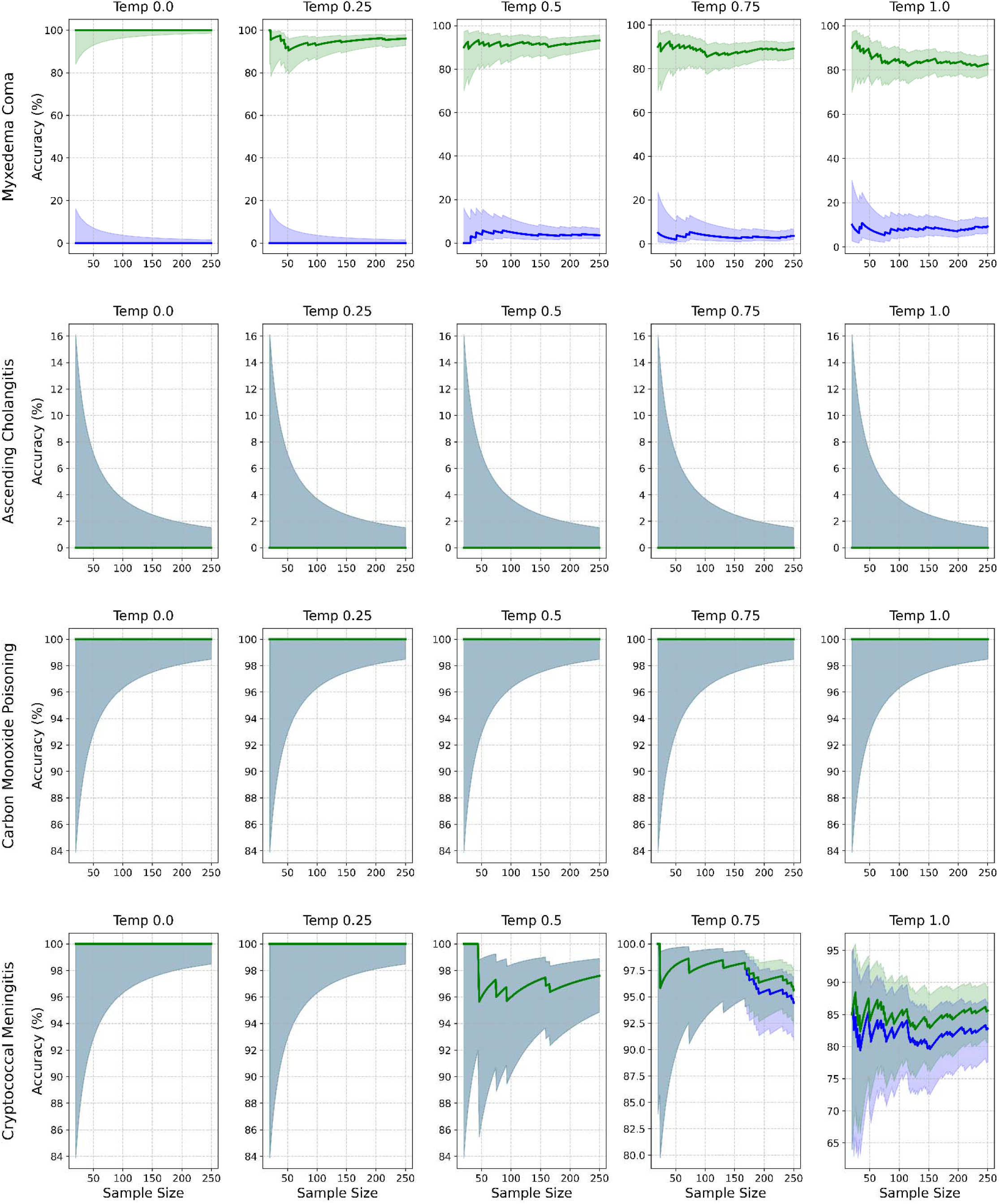
Diagnostic accuracy by case (rows) and temperature (columns) without exams. Lines show accuracy for leading (blue) and differential (green) diagnoses. Shaded pink and yellow areas represent the 95% confidence interval (CI) for the leading and differential diagnoses, respectively. Grey areas represent overlap in 95% CI.

**Supplement 3.**
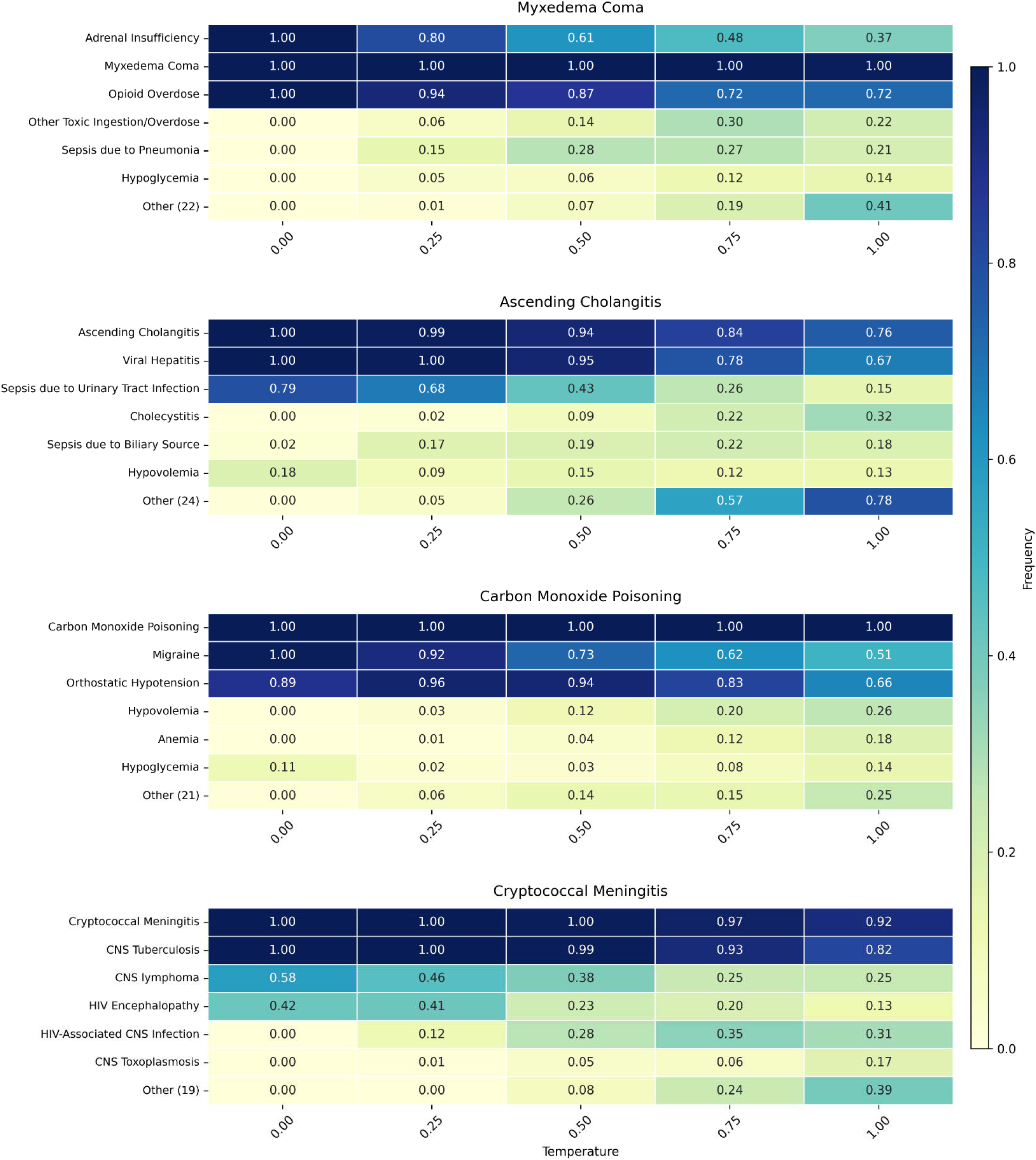
Heatmap depicting the top standardized diagnoses in the three-item differential with their corresponding frequency at each case-temperature pair. The “Other” diagnosis represents the cumulative frequency of all additional diagnoses considered for each case.

